# Statistical Analysis Plan for the SImPLE Trial: A Study to test if illustrations and plain language ImProve cLinical trial Education

**DOI:** 10.64898/2026.07.26.26358619

**Authors:** Stacey Llewellyn, Chris Gianacas, Craig Gedye, Emma Manifold

## Abstract

**Background:** Traditional participant information and consent forms (PICFs) have changed little over time and are often lengthy and complex. A novel consent process incorporating plain language, an infographic brochure and a short explanatory video was developed to improve communication of clinical trial information. The SImPLE study was designed to evaluate whether this novel consent process improves comprehension of clinical trial information, participant engagement and the consent experience compared with a traditional PICF.

**Design and Setting:** SImPLE is a randomised two-period crossover study involving adults receiving cancer treatment who are potentially eligible for clinical trial participation. Study materials were developed for the ANZadapt (ANZUP2101) prostate cancer clinical trial; however, participants are not being recruited to ANZadapt itself. Participants are randomised to receive either a standard 17-page NHMRC-style PICF followed by a novel consent process, or the reverse sequence, with a washout period of at least 7 days between study periods. The novel consent process comprises a simplified 4-page PICF, infographic brochure and short explanatory video. Twenty-four hours following each PICF review, participants complete a comprehension assessment and structured interview.

**Outcomes and Endpoints:** The primary endpoint is comprehension of clinical trial information, measured using a study-specific 22-item questionnaire comprising 10 true/false and 12 multiple-choice questions. Secondary endpoints include acceptability, assessed through participant preference for communication format, and engagement, assessed through confidence in explaining the trial and likelihood of agreeing to participate.

**Planned Analyses:** The primary analysis will compare comprehension scores between the novel and standard consent approaches using a paired t-test among participants completing both study periods. Secondary outcomes relating to acceptability and engagement will be summarised descriptively and compared between consent approaches as appropriate. Sensitivity analyses will be performed to assess the robustness of the primary findings and the potential impact of crossover design effects.

## 1 Administrative information

### 1.1 Study Identifiers

Protocol Date: 13 October 2025

Protocol: ANZUP2503 version1.1

Sponsor: ANZUP Cancer Trials Group Ltd

Level 8, 55 Botany Street

Randwick NSW 2031

Australia

### 1.2 Revision History

| Version | Date | Details |
| --- | --- | --- |
| 0.1 | 13July2026 | Initial draft by Stacey Llewellyn |
| 1.0 | 20July2026 | Final version |

### 1.3 Contributors to the statistical analysis plan

#### 1.3.1 Roles and Responsibilities

1.3 Contributors to the statistical analysis plan 1.3.1 Roles and Responsibilities
| Name | Affiliation | Role on study | SAP contribution |
| --- | --- | --- | --- |
| Stacey Llewellyn | The George Institute | Study statistician | Main author |
| Dr. Chris Gianacas | The George Institute | Senior statistician | Reviewed SAP |
| Dr. Craig Gedye | Icon Cancer Centre<br>Kurralta Park | Co-principal<br>investigator | Reviewed SAP |
| Dr. Emma Manifold | Icon Cancer Centre<br>Kurralta Park | Co-principal<br>investigator | Reviewed SAP |

#### 1.3.2 Approvals

The undersigned have reviewed this plan and approve it as final. They find it to be consistent with the requirements of the protocol as it applies to their respective areas.

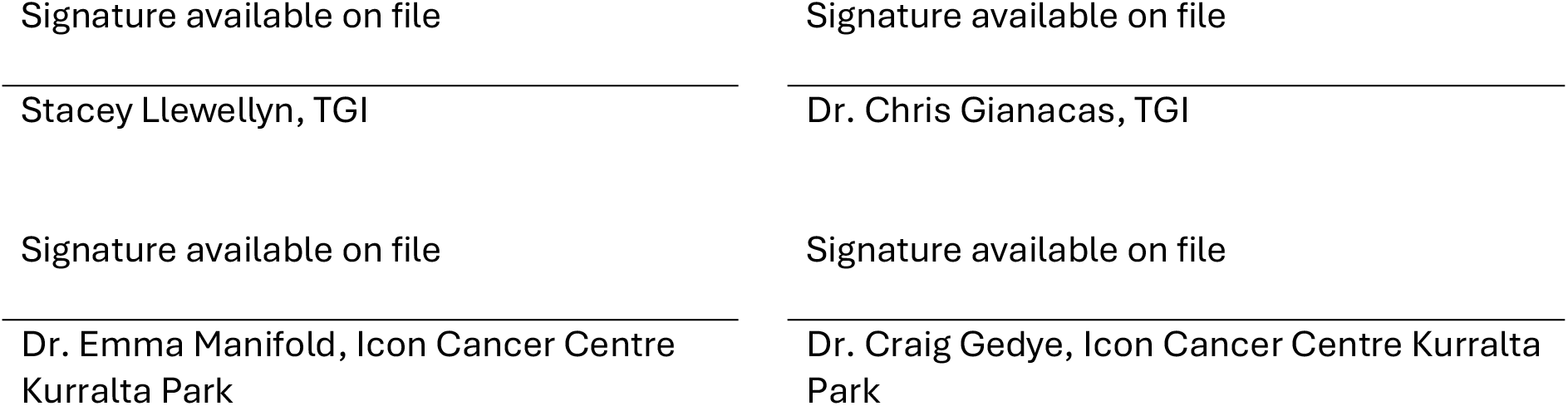

## 2 Study Overview

SImPLE is a randomised two-period crossover study designed to evaluate whether a novel multimedia participant information and consent process improves comprehension of clinical trial information compared with a traditional NHMRC-style participant information and consent form (PICF). The novel consent process consists of a simplified 4-page PICF, an infographic brochure and a short explanatory video, while the control consists of a standard 17-page PICF developed using the conventional NHMRC template. The study uses materials developed for the ANZadapt (ANZUP2101) prostate cancer clinical trial [1].

Participants are adults receiving cancer treatment who are eligible for participation in clinical trials but are not participants in the ANZadapt trial itself. Participants are randomised to one of two sequences:

- **Sequence 1:** Control (standard NHMRC PICF) followed by intervention (3-part communication package)
- **Sequence 2:** Intervention (3-part communication package) following by control (standard NHMRC PICF)

The crossover design was selected to allow each participant to act as their own control, thereby reducing between-participant variability and increasing statistical efficiency. A washout period of at least 7 days separates the two study periods. Twenty-four hours following each intervention, participants complete a quantitative comprehension assessment consisting of 10 true/false and 12 multiple-choice questions, together with a structured qualitative interview. At the end of the second study period, participants also complete questions relating to acceptability and consent preference.

The primary analysis will compare comprehension scores achieved under the novel and standard consent approaches using paired data from participants completing both study periods. Sensitivity analyses will assess the potential influence of period effects.

### 2.1 Sample Size

The primary outcome of this trial is the quality of informed consent, assessed quantitatively by a trial-specific instrument – an adaptation of the MICCA [2]. The primary endpoint is measured quantitatively with 10 true/false and 12 multiple-choice questions, with simple unweighted sum of up to 22 as the final score. A paired design will be used to test whether there is a paired mean difference.

A clinically meaningful difference would be 11/22 (50%) for the traditional PICF and 15/22 (68%) for the new consent process. A sample size of 30 participants would be required to detect this pared mean difference of 4 using a two-sided, paired-difference t-test; assuming a SD of the paired difference distribution of 6.6 (or less), 90% power and two-sided alpha of 0.05.

## 3 Study Aims and Objectives

### Aim

To test if a novel participant information and consent form improves comprehension of cancer clinical trial information compared with a traditional participant information and consent form.

### Hypothesis

1. A novel participant information and consent package is more comprehensible, efficient and acceptable than the traditional NHMRC PICF template.
2. A novel participant information and consent package elicits more engagement and interest in participation than the traditional NHMRC PICF template

### Primary objective

To quantitatively test comprehension of cancer clinical trial information comparing a novel versus a traditional participant information and consent form

### Secondary Objectives

1. To qualitatively test comprehension of cancer clinical trial information comparing a novel versus a traditional participant information and consent form*
2. To explore people’s preferences for these communication formats and identify opportunities to improve understanding clinical trial information.
3. To test if a particular format of information is more likely to engage and enlist people taking cancer treatment to take part in a clinical trial.

*Objective will not be covered in the is SAP

## 4 Study Endpoints

### 4.1 Primary Endpoint

Comprehension as measured quantitatively by the combined score of 10 true/false questions and 12 multiple-choice questions developed as a study-specific comprehension instrument via the MICCA/BIQ process. The maximum score that can be achieved is 22.

### 4.2 Secondary Endpoints

#### 4.2.1 Acceptability

Acceptability as defined by participants’ preference of which communication format is preferred between traditional vs. novel methods.

Acceptability will be assessed following completion of both consent methods by asking participants: “Which form of the trial communication did you prefer?”

Response options are:

- The Standard Consent Form.
- Brochure + Video + Short Consent Form.
- I did not prefer one or the other.
- I did not like either of the forms.

#### 4.2.2 Engagement

Engagement as defined by participants’ willingness to engage in or recommended the trial between traditional vs novel methods.

Engagement will be assessed using the following participant-reported measures collected following each consent method:

- Confidence in understanding the trial sufficiently to explain it to a friend or relative (5-level ordinal scale from “Very confident” to “Not confident at all”).
- Likelihood of agreeing to participate in the clinical trial based on the information provided (5-level ordinal scale from “Very likely” to “Not at all”).

## 5 Study Population

The study population briefly consists of adults over 18 years, eligible to participate in cancer clinical trials, but not asked to consent to the original trial itself. Full inclusion and exclusion criteria are provided in the study protocol.

### 5.1 Analysis population

The Analysis Population will include all participants who complete both study periods and have outcome data available from both assessments.

The analysis population used for each endpoint will be described in the relevant analysis section.

### 5.2 Subject Disposition

The flow of patients through this trial will be displayed in a CONSORT diagram. The report will include the number of participants who were assessed for eligibility, enrolled in the study, completed each period of the study (both intervention assessments) and were included in final analysis. Missing assessment by sequence will also be summarized.

### 5.3 Subject Characteristics

Subject characteristics will be summarized for the full analysis population overall and by randomisation group. Summaries will include:

- Demographics: Age, sex, language (English as first or second language (CALD)), highest educational level (tertiary, or secondary/below).
- Clinical characteristics: Tumour type and current treatment if any (i.e. chemotherapy, hormone therapy, immunotherapy)

Descriptive statistics will be used:

- Continuous variables: mean, standard deviation (SD), median and interquartile range (Q1-Q3), minimum, maximum
- Categorical variables: counts and percentages, with percentages calculated according to the number of participants for who data are available

## 6 General Statistical Considerations and Data Handling

### 6.1 General Principles

Analyses will be conducted primarily using SAS Enterprise Guide (version 9.3 or above) and R (version 4.3.1 or above). The significance level (alpha) will be set at 0.05, and all hypothesis tests will be two-sided, unless otherwise specified. Confidence intervals (CIs) will be reported at the 95% level.

### 6.2 Missing Data

Analyses will be based on available data. Imputation methods will not be considered for this study, however missingness will be described. Participants who have completed only one assessment will be included in descriptive summaries and sensitivity analyses as appropriate but will not contribute to the paired primary analysis.

## 7 Statistical Methods

### 7.1 Primary analysis

The primary analysis will compare comprehension scores between the novel consent process and the standard PICF using paired data from participants who complete both assessments (i.e. the analysis population). For each participant, the within-participant difference will be calculated as Novel score minus Standard PICF score. The mean within-participant difference, 95% confidence interval and p-value from a paired t-test will be reported.

Additionally, descriptive summaries including the mean, SD, median and interquartile range (IQR) will be reported for each of the comprehension scores by intervention, and intervention within study period.

### 7.2 Sensitivity analyses

Results from the following sensitivity analyses will be used to assess the robustness of conclusions to potential carryover effects. Sensitivity analyses will be considered supportive and exploratory rather than confirmatory.

#### 7.2.1 Wilcoxon signed-rank test

As a sensitivity analysis to the primary paired t-test, comprehension scores will be compared using a Wilcoxon signed-rank test for the analysis population. Median scores (IQR) for each assessment, the median paired difference (IQR), and the corresponding p-value will be reported.

#### 7.2.2 Treatment and period model

A linear mixed model will be fitted as a sensitivity analysis for the analysis population. The model will include fixed effects for treatment (Novel versus Standard) and period (Period 1 versus Period 2), together with a random participant effect to account for repeated measurements within individuals. Estimates, 95% confidence intervals and p-values for treatment and period effects will be reported. The estimated period effect will be used to assess the potential influence of the crossover-design effect on the primary analysis results [3]. Findings from this model will be interpreted as supportive evidence regarding the robustness of the treatment effect observed in the primary paired analysis.

#### 7.2.3 Period 1 Only Analysis

A sensitivity analysis restricted to Period 1 observations will also be performed for the analysis population. Participants randomised to the Standard followed by Novel sequence will contribute only their Standard PICF score, while participants randomised to the Novel followed by Standard sequence will contribute only their Novel consent process score. Comprehension scores will be compared between groups using a two-sample t-test. The mean difference, corresponding 95% confidence interval and p-value will be reported.

As participants have been exposed to only a single consent method at Period 1, this analysis may be less susceptible to period effects and will be interpreted as a supportive sensitivity analysis

### 7.3 Secondary analyses

#### 7.3.1 Acceptability

Acceptability will be assessed by participant preference for the standard consent form, novel consent process, neither, or no preference. Responses will be summarised using frequencies and percentages overall and by randomised sequence, with participants in the analysis population included. No formal hypothesis testing is planned.

#### 7.3.2 Engagement

Engagement will be assessed using two participant-reported measures collected following each consent method for the analysis population. Responses for each measure will be summarised separately using frequencies and percentages for each response category by consent method. Comparisons between consent methods will be performed using a Wilcoxon signed-rank test [4].

## Data Availability

No data are associated with this statistical analysis plan. Data reported in subsequent publications will be made available upon reasonable request, subject to applicable ethical and governance requirements.

## 9 Proposed Tables and Figures

**Figure 1.**
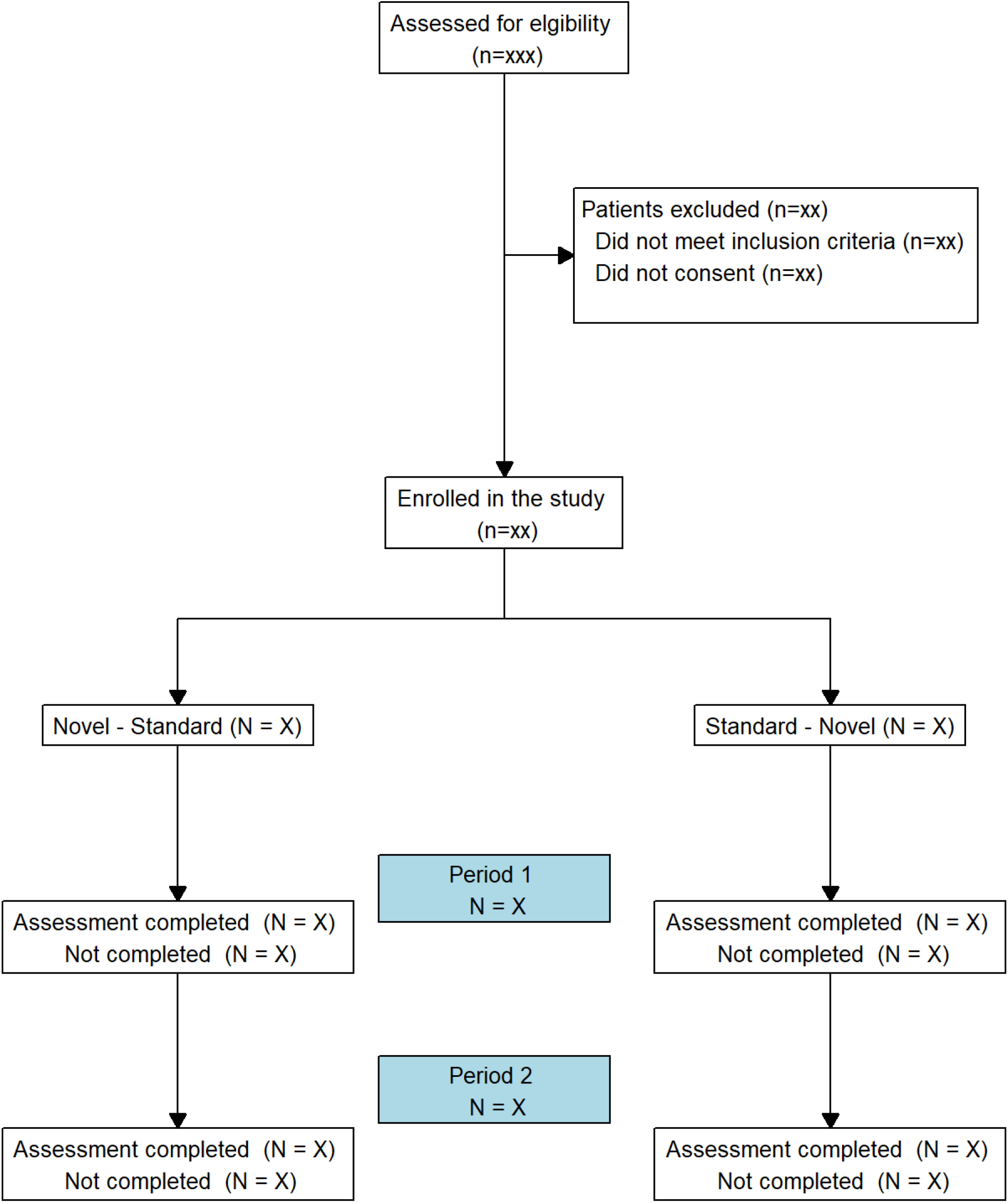
Consort flow diagram

**Table 1.** Participant characteristics.

|  | Sequence 1:<br>Standard - Novel<br>(N=XXX) | Sequence 2:<br>Novel - Standard<br>(N=XXX) | Overall<br>(N=XXX) |
| --- | --- | --- | --- |
| **Sex** |  |  |  |
| Male | xxx/xxx (xx.x%) | xxx/xxx (xx.x%) | xxx/xxx (xx.x%) |
| Female | xxx/xxx (xx.x%) | xxx/xxx (xx.x%) | xxx/xxx (xx.x%) |
| Other | xxx/xxx (xx.x%) | xxx/xxx (xx.x%) | xxx/xxx (xx.x%) |
| **Age (years)** |  |  |  |
| N | xxx | xxx | xxx |
| Mean (SD) | xx.x (xx.xx) | xx.x (xx.xx) | xx.x (xx.xx) |
| Median (IQR) | xx.x (xx.x; xx.x) | xx.x (xx.x; xx.x) | xx.x (xx.x; xx.x) |
| Min, Max | xx.x xx.x | xx.x xx.x | xx.x xx.x |
| **Language** |  |  |  |
| English as first language | xxx/xxx (xx.x%) | xxx/xxx (xx.x%) | xxx/xxx (xx.x%) |
| English as second language (CALD) | xxx/xxx (xx.x%) | xxx/xxx (xx.x%) | xxx/xxx (xx.x%) |
| **Highest educational level** |  |  |  |
| Tertiary education | xxx/xxx (xx.x%) | xxx/xxx (xx.x%) | xxx/xxx (xx.x%) |
| Secondary or below | xxx/xxx (xx.x%) | xxx/xxx (xx.x%) | xxx/xxx (xx.x%) |
| **Tumour Type** |  |  |  |
| XXX | xxx/xxx (xx.x%) | xxx/xxx (xx.x%) | xxx/xxx (xx.x%) |
| XXX | xxx/xxx (xx.x%) | xxx/xxx (xx.x%) | xxx/xxx (xx.x%) |
| XXX | xxx/xxx (xx.x%) | xxx/xxx (xx.x%) | xxx/xxx (xx.x%) |
| **Current Treatment** |  |  |  |
| None | xxx/xxx (xx.x%) | xxx/xxx (xx.x%) | xxx/xxx (xx.x%) |
| Chemotherapy | xxx/xxx (xx.x%) | xxx/xxx (xx.x%) | xxx/xxx (xx.x%) |
| Hormone therapy | xxx/xxx (xx.x%) | xxx/xxx (xx.x%) | xxx/xxx (xx.x%) |
| Immunotherapy | xxx/xxx (xx.x%) | xxx/xxx (xx.x%) | xxx/xxx (xx.x%) |

**Table 2.** Comprehension descriptive summaries.

|  | N | Mean (SD) | Median (IQR) | Min, Max |
| --- | --- | --- | --- | --- |
| **Consent Method** |  |  |  |  |
| Standard PICF | xx | xx.x (xx.x) | xx.x (xx.x – xx.x) | x.xx – x.xx |
| Novel Consent | xx | xx.x (xx.x) | xx.x (xx.x – xx.x) | x.xx – x.xx |
| **Consent Method by Period** |  |  |  |  |
| Standard PICF, Period 1 | xx | xx.x (xx.x) | xx.x (xx.x – xx.x) | x.xx – x.xx |
| Standard PICF, Period 2 | xx | xx.x (xx.x) | xx.x (xx.x – xx.x) | x.xx – x.xx |
| Novel Consent, Period 1 | xx | xx.x (xx.x) | xx.x (xx.x – xx.x) | x.xx – x.xx |
| Novel Consent, Period 2 | xx | xx.x (xx.x) | xx.x (xx.x – xx.x) | x.xx – x.xx |

**Table 3.** Summary of primary outcome: comprehension.

|  | Standard PICF<br>Mean (SD)<br>(N=XXX) | Novel PICF<br>Mean (SD)<br>(N=XXX) | Paired Difference (95% CI)<br>(Novel-Standard)<br>(N=XXX) | p-value |
| --- | --- | --- | --- | --- |
| Comprehension<br>Score | xx.x (xx.x) | xx.x (xx.x) | xx.x (95% CI xx.x- xx.x) | x.xx |

**Table 4.** Summary of primary outcome sensitivity analyses: comprehension.

| Analysis | Estimate | 95% CI | p-value |
| --- | --- | --- | --- |
| **Wilcoxon signed-rank test** |  |  |  |
| Standard PICF, Median (IQR) | xx.x (xx.x – xx.x) | - | - |
| Novel PICF, Median (IQR) | xx.x (xx.x – xx.x) | - | - |
| Median paired difference (IQR)<br>(Novel-standard) | xx.x (xx.x – xx.x) | - | x.xx |
| **Linear mixed model** |  |  |  |
| Treatment effect | xx.x | xx.x – xx.x | x.xx |
| Period effect | xx.x | xx.x – xx.x | x.xx |
| **Period 1 only analysis** |  |  |  |
| Standard PICF, Mean (SD) | xx.x (xx.x) | - | - |
| Novel PICF, Mean (SD) | xx.x (xx.x) | - | - |
| Mean difference (Novel – Standard) | xx.x | xx.x – xx.x | x.xx |

**Table 5.** Summary of secondary outcomes assessing acceptability.

|  | Sequence 1:<br>Standard - Novel<br>(N=XXX) | Sequence 2:<br>Novel - Standard<br>(N=XXX) | Overall<br>(N=XXX) |
| --- | --- | --- | --- |
| **Which form of the trial communication did you prefer?** |  |  |  |
| The Standard Consent Form | xxx/xxx (xx.x%) | xxx/xxx (xx.x%) | xxx/xxx (xx.x%) |
| Brochure + Video + Short Consent Form | xxx/xxx (xx.x%) | xxx/xxx (xx.x%) | xxx/xxx (xx.x%) |
| I did not prefer one or the other | xxx/xxx (xx.x%) | xxx/xxx (xx.x%) | xxx/xxx (xx.x%) |
| I did not like either of the forms | xxx/xxx (xx.x%) | xxx/xxx (xx.x%) | xxx/xxx (xx.x%) |

**Table 6.** Summary of secondary outcomes assessing engagement.

|  | Standard PICF<br>(N=XXX) | Novel PICF<br>(N=XXX) | p-value<sup>^</sup> |
| --- | --- | --- | --- |
| **Confidence in understanding the trial** |  |  | X.XXX |
| Very confident | xxx/xxx (xx.x%) | xxx/xxx (xx.x%) |  |
| Somewhat | xxx/xxx (xx.x%) | xxx/xxx (xx.x%) |  |
| Unsure | xxx/xxx (xx.x%) | xxx/xxx (xx.x%) |  |
| Not confident | xxx/xxx (xx.x%) | xxx/xxx (xx.x%) |  |
| Not confident at all | xxx/xxx (xx.x%) | xxx/xxx (xx.x%) |  |
| **Likelihood of agreeing to participate in the trial** |  |  | X.XXX |
| Very likely | xxx/xxx (xx.x%) | xxx/xxx (xx.x%) |  |
| Possibly | xxx/xxx (xx.x%) | xxx/xxx (xx.x%) |  |
| Unsure | xxx/xxx (xx.x%) | xxx/xxx (xx.x%) |  |
| Unlikely | xxx/xxx (xx.x%) | xxx/xxx (xx.x%) |  |
| Not at all | xxx/xxx (xx.x%) | xxx/xxx (xx.x%) |  |
| <sup>^</sup>P-value obtained from a Wilcoxon signed-rank test comparing paired participant responses under the Novel and Standard consent methods. |  |  |  |

## Appendix

**Table 7.** Summary of correct responses to multiple-choice and true/false questions by PICF.

**Appendix***Table 7 Summary of correct responses to multiple-choice and true/false questions by PICF*
| Proportion correct (%) | Standard PICF<br>(N=XXX) | Novel PICF<br>(N=XXX) |
| --- | --- | --- |
| **Multiple choice questions** |  |  |
| Q1 | xxx/xxx (xx.x%) | xxx/xxx (xx.x%) |
| Q2 | xxx/xxx (xx.x%) | xxx/xxx (xx.x%) |
| Q3 | xxx/xxx (xx.x%) | xxx/xxx (xx.x%) |
| Q4 | xxx/xxx (xx.x%) | xxx/xxx (xx.x%) |
| Q6 | xxx/xxx (xx.x%) | xxx/xxx (xx.x%) |
| Q7 | xxx/xxx (xx.x%) | xxx/xxx (xx.x%) |
| Q8 | xxx/xxx (xx.x%) | xxx/xxx (xx.x%) |
| Q9 | xxx/xxx (xx.x%) | xxx/xxx (xx.x%) |
| Q10 | xxx/xxx (xx.x%) | xxx/xxx (xx.x%) |
| Q11 | xxx/xxx (xx.x%) | xxx/xxx (xx.x%) |
| Q12 | xxx/xxx (xx.x%) | xxx/xxx (xx.x%) |
| **True-False questions** |  |  |
| Q1 | xxx/xxx (xx.x%) | xxx/xxx (xx.x%) |
| Q2 | xxx/xxx (xx.x%) | xxx/xxx (xx.x%) |
| Q3 | xxx/xxx (xx.x%) | xxx/xxx (xx.x%) |
| Q4 | xxx/xxx (xx.x%) | xxx/xxx (xx.x%) |
| Q6 | xxx/xxx (xx.x%) | xxx/xxx (xx.x%) |
| Q7 | xxx/xxx (xx.x%) | xxx/xxx (xx.x%) |
| Q8 | xxx/xxx (xx.x%) | xxx/xxx (xx.x%) |
| Q9 | xxx/xxx (xx.x%) | xxx/xxx (xx.x%) |
| Q10 | xxx/xxx (xx.x%) | xxx/xxx (xx.x%) |

**Table 8.** Summary of correct responses to multiple-choice and true/false questions by PICF and study period.

| Proportion correct (%) | Period 1 |  | Period 2 |  |
| --- | --- | --- | --- | --- |
|  | Standard PICF<br>(N=XXX) | Novel PICF<br>(N=XXX) | Standard PICF<br>(N=XXX) | Novel PICF<br>(N=XXX) |
| **Multiple choice questions** |  |  |  |  |
| Q1 | xxx/xxx (xx.x%) | xxx/xxx (xx.x%) | xxx/xxx (xx.x%) | xxx/xxx (xx.x%) |
| Q2 | xxx/xxx (xx.x%) | xxx/xxx (xx.x%) | xxx/xxx (xx.x%) | xxx/xxx (xx.x%) |
| Q3 | xxx/xxx (xx.x%) | xxx/xxx (xx.x%) | xxx/xxx (xx.x%) | xxx/xxx (xx.x%) |
| Q4 | xxx/xxx (xx.x%) | xxx/xxx (xx.x%) | xxx/xxx (xx.x%) | xxx/xxx (xx.x%) |
| Q6 | xxx/xxx (xx.x%) | xxx/xxx (xx.x%) | xxx/xxx (xx.x%) | xxx/xxx (xx.x%) |
| Q7 | xxx/xxx (xx.x%) | xxx/xxx (xx.x%) | xxx/xxx (xx.x%) | xxx/xxx (xx.x%) |
| Q8 | xxx/xxx (xx.x%) | xxx/xxx (xx.x%) | xxx/xxx (xx.x%) | xxx/xxx (xx.x%) |
| Q9 | xxx/xxx (xx.x%) | xxx/xxx (xx.x%) | xxx/xxx (xx.x%) | xxx/xxx (xx.x%) |
| Q10 | xxx/xxx (xx.x%) | xxx/xxx (xx.x%) | xxx/xxx (xx.x%) | xxx/xxx (xx.x%) |
| Q11 | xxx/xxx (xx.x%) | xxx/xxx (xx.x%) | xxx/xxx (xx.x%) | xxx/xxx (xx.x%) |
| Q12 | xxx/xxx (xx.x%) | xxx/xxx (xx.x%) | xxx/xxx (xx.x%) | xxx/xxx (xx.x%) |
| **True-False questions** |  |  |  |  |
|  | Standard PICF<br>(N=XXX) | Novel PICF<br>(N=XXX) | Standard PICF<br>(N=XXX) | Novel PICF<br>(N=XXX) |
| Q1 | xxx/xxx (xx.x%) | xxx/xxx (xx.x%) | xxx/xxx (xx.x%) | xxx/xxx (xx.x%) |
| Q2 | xxx/xxx (xx.x%) | xxx/xxx (xx.x%) | xxx/xxx (xx.x%) | xxx/xxx (xx.x%) |
| Q3 | xxx/xxx (xx.x%) | xxx/xxx (xx.x%) | xxx/xxx (xx.x%) | xxx/xxx (xx.x%) |
| Q4 | xxx/xxx (xx.x%) | xxx/xxx (xx.x%) | xxx/xxx (xx.x%) | xxx/xxx (xx.x%) |
| Q6 | xxx/xxx (xx.x%) | xxx/xxx (xx.x%) | xxx/xxx (xx.x%) | xxx/xxx (xx.x%) |
| Q7 | xxx/xxx (xx.x%) | xxx/xxx (xx.x%) | xxx/xxx (xx.x%) | xxx/xxx (xx.x%) |
| Q8 | xxx/xxx (xx.x%) | xxx/xxx (xx.x%) | xxx/xxx (xx.x%) | xxx/xxx (xx.x%) |
| Q9 | xxx/xxx (xx.x%) | xxx/xxx (xx.x%) | xxx/xxx (xx.x%) | xxx/xxx (xx.x%) |
| Q10 | xxx/xxx (xx.x%) | xxx/xxx (xx.x%) | xxx/xxx (xx.x%) | xxx/xxx (xx.x%) |

